# Plasma Microbial Cell-Free DNA Metagenomic Sequencing Bridges Gaps in the Diagnosis, Epidemiology and Surveillance of *Legionella* Infections

**DOI:** 10.64898/2026.03.23.26348694

**Authors:** Frederick S. Nolte, Martin S. Lindner, Shivkumar Venkatasubrahmanyam, Chiagozie I. Pickens, Lindsay Lim, Vincent P. Hsu, Sarah Y. Park, Bradley A. Perkins, Richard. G Wunderink

**Affiliations:** Karius, Inc., Redwood City, CA; Northwestern University Feinberg School of Medicine, Chicago, IL; Loma Linda University School of Medicine and AdventHealth, Orlando, FL and; University of Central Florida College of Medicine and AdventHealth, Orlando, FL

## Abstract

Conventional diagnostic methods (CDM) for *Legionella* preferentially detect *L. pneumophila* and frequently fail to identify non-pneumophila species (NPLS), obscuring the full clinical spectrum of infection and limiting surveillance accuracy. We analyzed plasma microbial cell-free DNA (mcfDNA) sequencing detections of *Legionella* spp. from a large clinical cohort tested between 2018 and 2024 and compared species distributions with culture and PCR confirmed cases reported in the most recent national surveillance datasets (2018-2021). To contextualize the clinical impact, we reviewed published reports in which mcfDNA sequencing was used to diagnose legionellosis (2021–2025) and evaluated real-world performance data from a hospital contributing 8.9% of detections within the cohort (Hospital A). mcfDNA sequencing identified proportionally fewer *L. pneumophila*, more NPLS, and fewer unresolved species than the CDC reports (all p<0.001). Among 15 publications describing 19 U.S. patients, 74% were immunocompromised and 79% had NPLS infections. Concordance between mcfDNA and CDM occurred in 31.6% of cases. At Hospital A with 36 detections, diagnosis was established by CDM alone in none, by both CDM and mcfDNA in 23.5%, and by mcfDNA alone in 76.5%, yielding an additive diagnostic value of 56.8% These findings suggest that plasma mcfDNA sequencing may improve detection of NPLS particularly in high-risk or diagnostically challenging patients and provide complementary data for both clinical diagnosis and epidemiologic surveillance.

**IMPORTANCE:** *Legionella* infections remain underdiagnosed due to heavy reliance on the urinary antigen test, which detects only *L. pneumophila* serogroup 1 as the primary diagnostic modality. Although culture and PCR have the potential to detect all *Legionella* spp., they are not widely implemented in clinical laboratories and may also fail to detect non-*pneumophila* species.

Plasma microbial cell-free DNA (mcfDNA) sequencing overcomes these limitations by providing broad, unbiased detection across the *Legionella* genus. Using a large, multi-year clinical cohort, we show that mcfDNA sequencing identifies more non-*pneumophila* species than captured in national surveillance data, revealing hidden patterns of disease and offering new insight into *Legionella* epidemiology. These findings highlight a major systemic gap between true clinical burden and reported cases. By detecting infections missed by conventional methods, plasma mcfDNA sequencing has the potential to improve patient outcomes. At the public health level, aggregated mcfDNA data represent a complementary surveillance resource capable of strengthening national monitoring efforts.

## INTRODUCTION

Legionnaires’ disease (LD) and Pontiac fever are illnesses caused by *Legionella* spp., which are commonly found in both natural and man-made aquatic environments. Transmission occurs through inhalation of aerosolized water droplets containing bacteria. Legionnaires’ disease is a severe form of pneumonia requiring prompt antibiotic therapy, whereas Pontiac fever is a self-limited, influenza-like illness that typically resolves without specific treatment. In addition to pulmonary disease, extrapulmonary *Legionella* infections can occur. These systemic infections, often seen in immunocompromised patients, result from hematogenous dissemination and may involve sites such as the skin, heart, or central nervous system (1).

In general, reported cases of LD in the US have been increasing since the early 2000s, with a peak in 2018 (1). The World Health Organization reported the overall mortality by LD as 5-10% and as high as 40-80% in immunosuppressed individuals (2).

Since LD cannot be reliably distinguished from other forms of pneumonia by clinical, radiographic or nonspecific laboratory studies, *Legionella* specific tests are necessary for diagnosis (3). Despite decades of surveillance, the epidemiology of *Legionella* infections remains poorly defined because conventional diagnostic methods (CDM) systematically under-detect non-*pneumophila* species, particularly in immunocompromised hosts.

Of the 66 validly named *Legionella* species, 26 are thought to be pathogenic for humans (4). *Legionella pneumophila* is typically recognized as the primary cause of LD. Although urinary antigen tests (UAT) are the main diagnostic tools in most countries, offering rapid results, they are limited by detecting only *L. pneumophila* serogroup 1 (Lp1). Culture, although able to detect all species and serogroups, has low sensitivity and requires inoculation of multiple selective media, extended incubation, and isolate identification. Consequently, culture is not widely deployed in clinical laboratories. Additionally, due to infrequent sputum production in LD patients, bronchoalveolar lavage (BAL) is often necessary to obtain samples. MALDI-TOF MS can provide species-level identification of *Legionella* from culture isolates and has high overall accuracy if the species is included in the database (5). Most systems now accurately identify the most common *Legionella* spp. but cannot identify all species.

Molecular diagnostics are more sensitive than culture. While numerous laboratory-developed tests exist, only two FDA-cleared, nucleic acid amplification tests for detection of *L. pneumophila* alone t are included as part of respiratory syndromic panels (BioFire Pneumonia Panel, bioMérieux and the Unyvero LRT Panel, Curetis). Multiplex assays are preferred since they detect both *L. pneumophila* and non-*pneumophila Legionella* species (NPLS); however, they do not differentiate between individual NPLS (6).

Broad range PCR with sequencing of 16s RNA gene amplicons can be used to identify colonies and direct detection of *Legionella* spp. in tissue and body fluid samples. While partial sequencing of the 16S rRNA gene can accurately identify all *Legionella* spp. to the genus level and all *L. pneumophila* serogroups, it often leads to incorrect species-level identification for some of the less common NPLS (4).

Unbiased plasma microbial cell free DNA (mcfDNA) sequencing [Karius Spectrum^TM^ (KS)] reports mcfDNA detections and concentrations in molecules/μl (MPM) for >1,000 human pathogens including 29 *Legionella* spp. for non-invasive diagnosis of infections throughout the body (7). It is available a referral test through Karius’s CLIA licensed, CAP accredited clinical laboratory in Redwood City, CA currently used by >500 healthcare systems in the U.S.

A previous analysis of 18,690 reports from 15,165 patients processed using the KS test identified 80 reports of 13 different *Legionella* species highlighting its potential as a pan *Legionella* diagnostic (8). A phylogeny-based filter for mapping and reporting reads as a method for identifying species (DC-3.16) was recently incorporated into the current validated commercial pipeline (9). It increases the confidence in species identification by addressing pathogen cross-reactivity, limited representation within the genome database and unresolved species.

To address the gaps in CDM, we investigated the potential of plasma mcfDNA sequencing as a pan *Legionella* diagnostic tool. This study characterizes *Legionella* spp. detections identified by plasma mcfDNA sequencing in a larger cohort derived from a commercial testing database. To contextualize these findings, we compared species distribution with culture and PCR-confirmed cases reported in the U.S. national surveillance data from 2018 to 2021 (10, 11) and reviewed all published case reports in which plasma mcfDNA sequencing was used to diagnose legionellosis. Finally, we present real-world evidence of test performance and clinical utility from a hospital contributing a substantial number of *Legionella* detections within this cohort.

## MATERIAL AND METHODS

### KS cohort

Plasma mcfDNA metagenomic sequencing was performed as previously described (7). All pre-DC-3.16 production results with ≥3 legionellae DNA fragments out of 78,527 tests from > 400 hospitals from 2018-2024 were reanalyzed with DC-3.16. *Legionella* reads in 15,518 no template controls (NTC) and in a 684 asymptomatic patient cohort were also analyzed with DC-3.16 to address concerns about environmental contamination and colonization with legionellae, respectively. Patient information obtained from test request forms (TRF) including age, sex, originating laboratory city and state, ICD-10 codes and any other indicators of underlying patient conditions were reviewed to provide clinical context for orders and geographic distribution. KS results for those patients who had *Legionella* spp. detected in multiple samples from the same patient were examined to investigate the reproducibility of the detections and the duration and kinetics of the mcfDNA signal in plasma.

### CDC cohort

The relative frequencies of detection of the varied species within the KS cohort were compared to those confirmed by culture and PCR, as reported by the CDC. This comparison utilized data from the two most recent Legionnaires’ Disease Surveillance Summary Report, which summarized reports for 2018 and 2019 (10) and 2020 and 2021 (11). PCR for case confirmation was reported only in the latter report.

### Literature review

To inform the correlation KS detections with CDM and potential additive diagnostic value, all publications in which KS was used to diagnose legionellosis were reviewed. Publications were drawn from the Karius clinical evidence library and supplemented by a PubMed search using the search terms microbial cell-free DNA, Karius, and *Legionella*.

### Hospital A cohort

A data use agreement was established with the healthcare system that had 36 (9.6%) the total *Legionella* detections in the KS Cohort. The following data were collected for these patients: age, gender, illness onset date, hospital admission date, acquisition, clinical presentation, risk factors, comorbidities, outcome, results of all CDM performed and clinical impact. Clinical impact definitions included earlier, new or confirmed diagnoses and change in therapy.

### Statistical analysis

A Chi-squared test for the comparison of two proportions from independent samples was used to determine significance levels between species identified in the two cohorts (12).

### Ethical considerations

Under Advarra IRB KAR-0028, Use of Leftover Clinical Diagnostic Samples for Research and Development of Karius Metagenomic Sequencing Assays and Product Development (UMBRELLA) (Pro00087833) covered the access of PHI and was waived from oversite. Under the data use agreement with Hospital A the Data User only disclosed the PHI received under this Data Use Agreement for the purpose of performing the Research. As a condition of receiving the Limited Data Set, Data User agreed to protect the privacy of the Limited Data Set in accordance with the terms and conditions of the Agreement and all applicable federal and state privacy and security laws, including but not limited to HIPAA and the HIPAA Regulations.

## RESULTS

### KS cohort

Examination of the raw data for all production results during the study period revealed 610 (0.78%) samples with ≥3 Legionella mcfDNA fragments. Reanalysis of these samples with DC-3.16 produced 404 *Legionella* calls that passed all statistical criteria for reporting from 376 unique patients. These calls represented 14 different taxa with a median concentration of 1,781 MPM (IQR 262-24,025). The calls were 171 (42.2%) *L. pneumophila,* 201 (49.8%) NPLS and 32 (7.9%) unresolved *Legionella* sp. (Fig.1). The numbers of the individual NPLS are shown in Fig. 2. A single species was reported for each sample, except one in which both *L. pneumophila* and *L. dumoffii* were reported (Supplemental Fig 1A). *Legionella* spp. were detected by KS in multiple samples (from 2 to 5) from 20 patients, all with the same species: 6, *pneumophila*; 4 unresolved species, 3 *micdadei*; 2, *bozemanae*; 2, *maceachernii*; 2, *longbeachae* and 1, *jordanis* (Supplemental Table 1). Median duration of mcfDNA signal was 24.5 days (IQR 12.5-55.8). Signal decreased between first and last samples in 17 patients (median -2.5 log, IQR -3.26 to -0.70), increased in two (0.89 and 4.4 log) and remained the same in one at 3.3 log.

**Figure 1.**
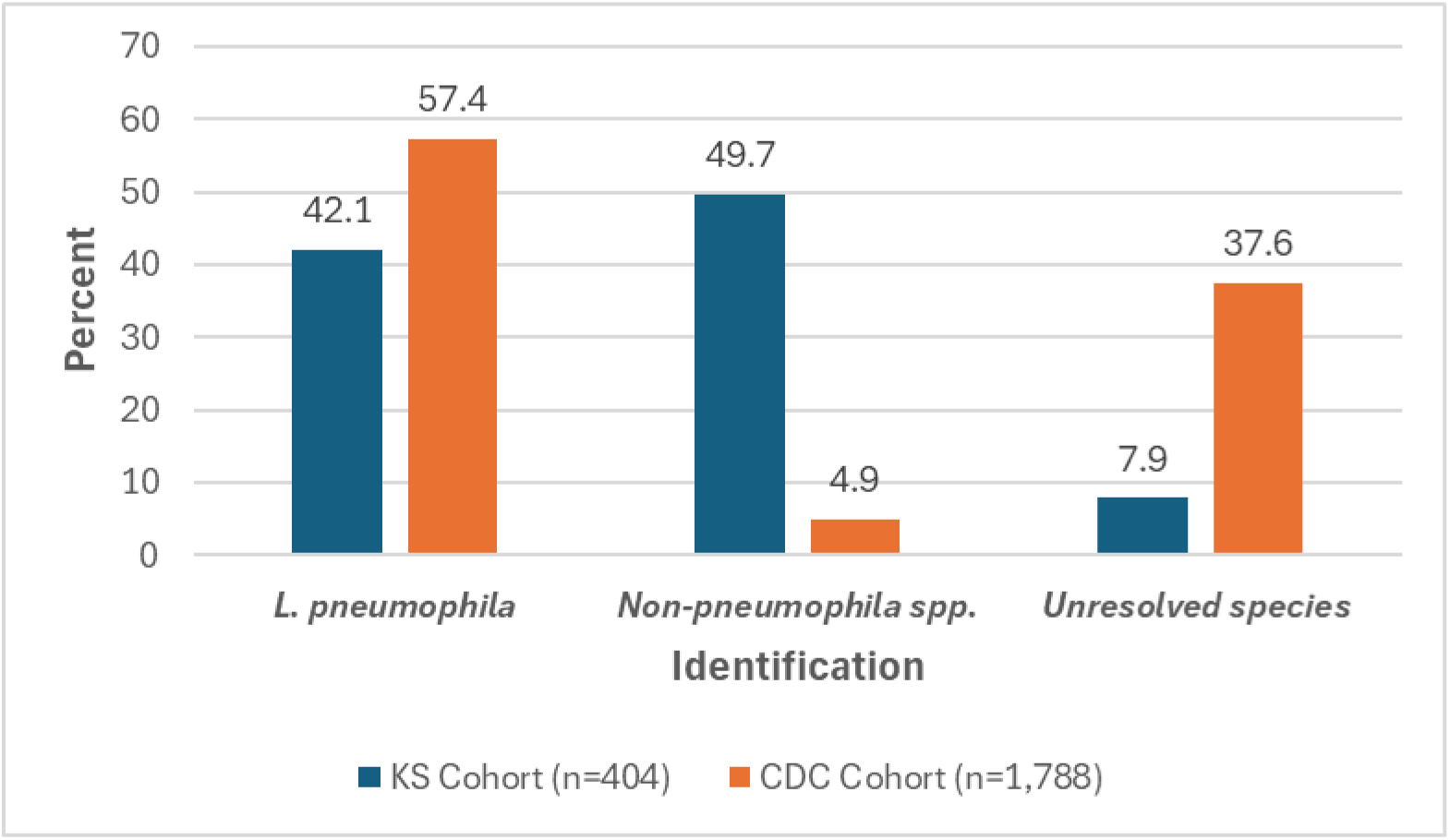
Comparisons of the proportions of *L. pneumophila*, non-*pneumophila* species and unresolved species reported in the Karius Spectrum and CDC cohorts. All differences were statistically significant (p<0.001).

**Figure 2.**
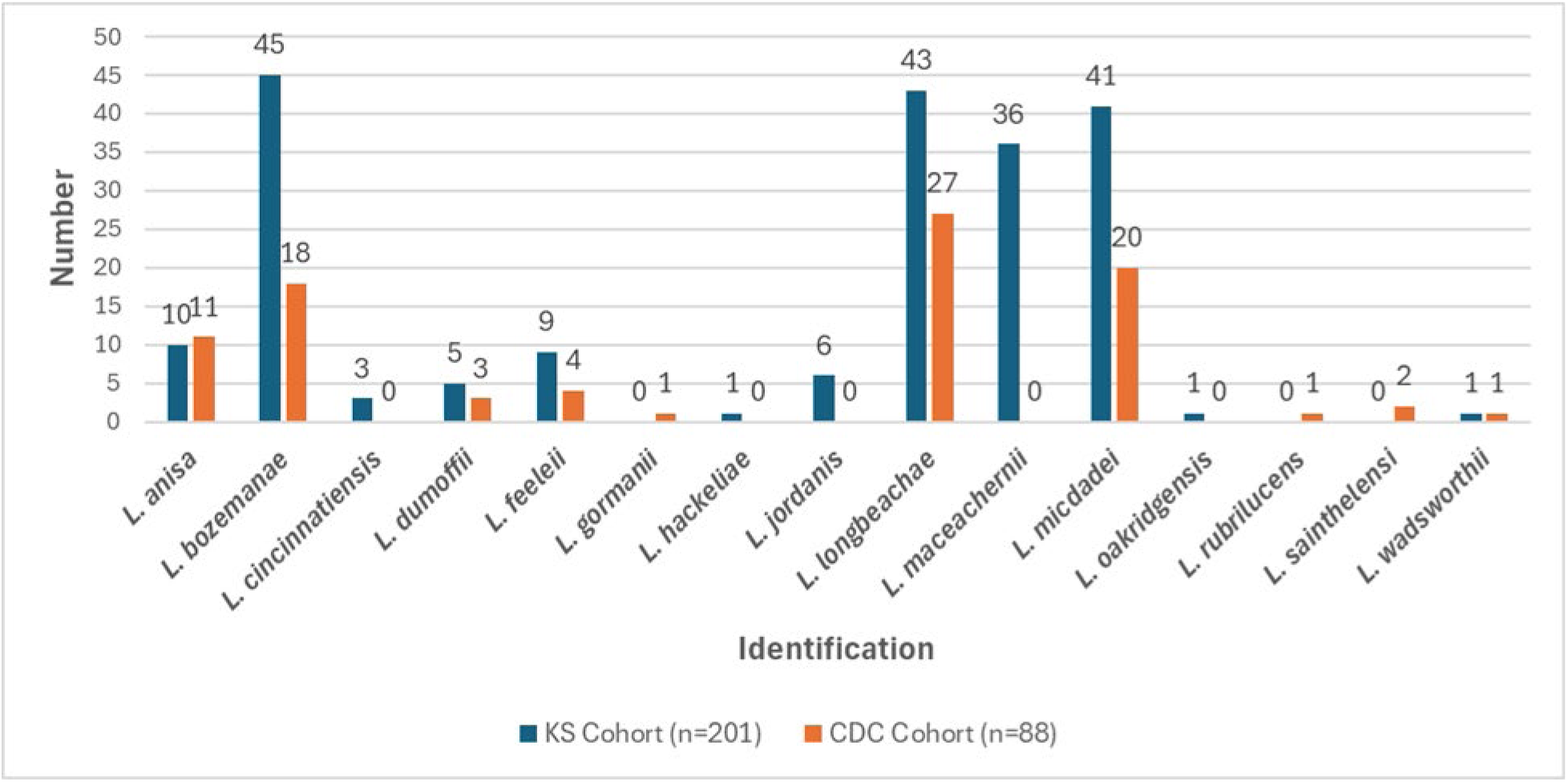
Number of non-*pneumophila* species identified in the Karius Spectrum and CDC cohorts.

For the 376 patients for which a *Legionella* sp. was reported, 61.7% were male and 88.4% were ≥18 years old. ICD-10 codes provided on 37.1% of TRF represented a diverse array of clinical scenarios, with 66.9% consistent with pulmonary manifestations and 21.2% indicating an immunocompromising condition. Samples were obtained from hospitalized patients from referring laboratories in 30 states. A heat map of the number of KS *Legionella* spp. reports by state are shown in Fig. 3. A review of detections by referring institutions revealed no temporal clustering of individual species suggestive of an outbreak.

**Figure 3.**
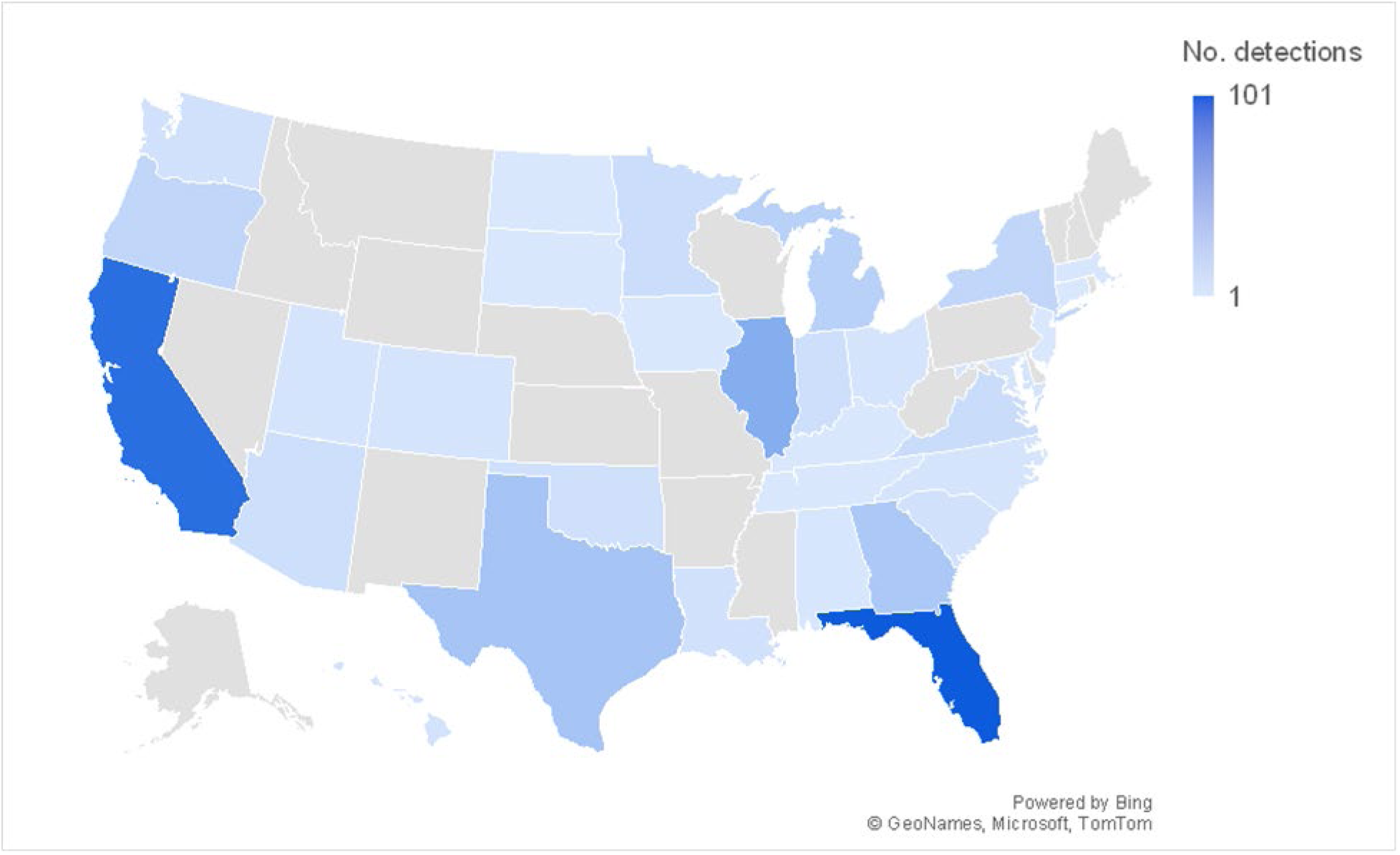
Heat map of plasma mcfDNA sequencing detections of *Legionella* species by state of the referring healthcare facility.

Only 0.02% of the NTC had ≥5 reads, all *L. pneumophila*, which is analytically necessary but not sufficient for reporting due to failure to meet all statistical criteria. Analysis of plasma from the 687 asymptomatic donors showed background levels for *L. maceachernii*, which would have been reported twice (0.3%) and a small number of reads (median 2, range 1-9) were observed in 78 (12.6%) others. Other *Legionella* species were present, but the number of reads was extremely low (median 1, range 1-5), below the threshold for reporting.

### CDC cohort

The number of reported confirmed cases of LD (31,128) by diagnostic testing method were 29342 (94.3%) UAT, 1306 (4.2%) culture, 481 (1.5%) PCR and 12 (0.04%) serology. We chose culture and PCR as the comparators since they have the potential to detect all *Legionella* spp. The results reported to CDC by both methods included 1027 (67.7%) *L. pneumophila,* 88 (4.9%) NPLS and 673 (37.6%) unresolved *Legionella* sp. (Fig. 1). PCR contributed 206 (20.1%) *L pneumophila*, 6 (6.8%) NPLS and 296 (44%) unresolved species detections to the cohort. Geographic distribution, patient demographics, hospitalizations, outcomes, and type of legionellosis in the cases included in this cohort were previously described.

### Cohort comparisons

KS identified proportionally less *L. pneumophila*, more NPLS and less unresolved species than CDC culture reports (all p<0.001). There were 201 reports of NPLS in the KS cohort and 88 in the CDC cohort comprising 15 distinct species with 12 reported in KS and 10 in the CDC cohort. Five were identified in the KS cohort alone: 3, *L. cincinnatiensis*; 1, *L. hackeliae*; 6, *L. jordanis*; 36, *L. maceachernii*; and 1, *L. oakridgensis*. Three were identified in the CDC cohort alone: 1, *L. gormanii*; 2, *L. sainthelensi* and 1, *L. rubrilucens* (Fig.2). Although patient demographics were not stratified by confirmation method in the CDC cohort most cases occurred in patients ≥ 50 years of age like KS cohort. However, the KS cohort included a higher proportion of children (4.4% vs. 0.12%) and adolescents (8.2% vs. 0.17%); p<0.001 for both age groups. The percentage of males in both cohorts were also similar. Nearly all patients in both cohorts were admitted to hospital. In the CDC cohort the reported LD incidence was highest in the East North Central, Middle Atlantic, and New England regions. Because KS detections reflected regional variation in test utilization, the states with the highest numbers of KS detections differed from those with the highest reported incidence. Because the CDC and KS cohorts covered different time periods, we analyzed KS detections from 2022–2024, the years not included in CDC reporting, and compared them with the full KS cohort. No significant differences were observed in the relative proportions of *L. pneumophila*, NLPS, or unresolved species, and the spectrum of NLPS detected was similar (data not shown).

### Literature review

The literature review yielded 15 publications with 19 patients that reported the use of KS in the evaluation of patients with legionellosis and summarized in Table 1 (13–27). Patient characteristics included 84% adults, 58.8% male, 74% immunocompromised and 11% with extrapulmonary manifestations (suspected meningitis). Infections included 21% with *L. pneumophila* and 79% with 7 other species 4, *micdadei*; 4, *bozemanae*; 2, *hackeliae*; 2, *longbeachae*, 1 each *anisa*, *feeleii*, and *maceachernii*). Median concentration was 6,723 MPM (IQR 909-20,094). Invasive procedures were used to collect diagnostic specimens in 11 (58%) patients. BAL in 8, biopsy in 2 and both in 1. All patients were treated with antibiotics effective against legionellae, and of 14 patients whose outcomes were reported 13 (92.8%) survived their infection.

**Table 1.**
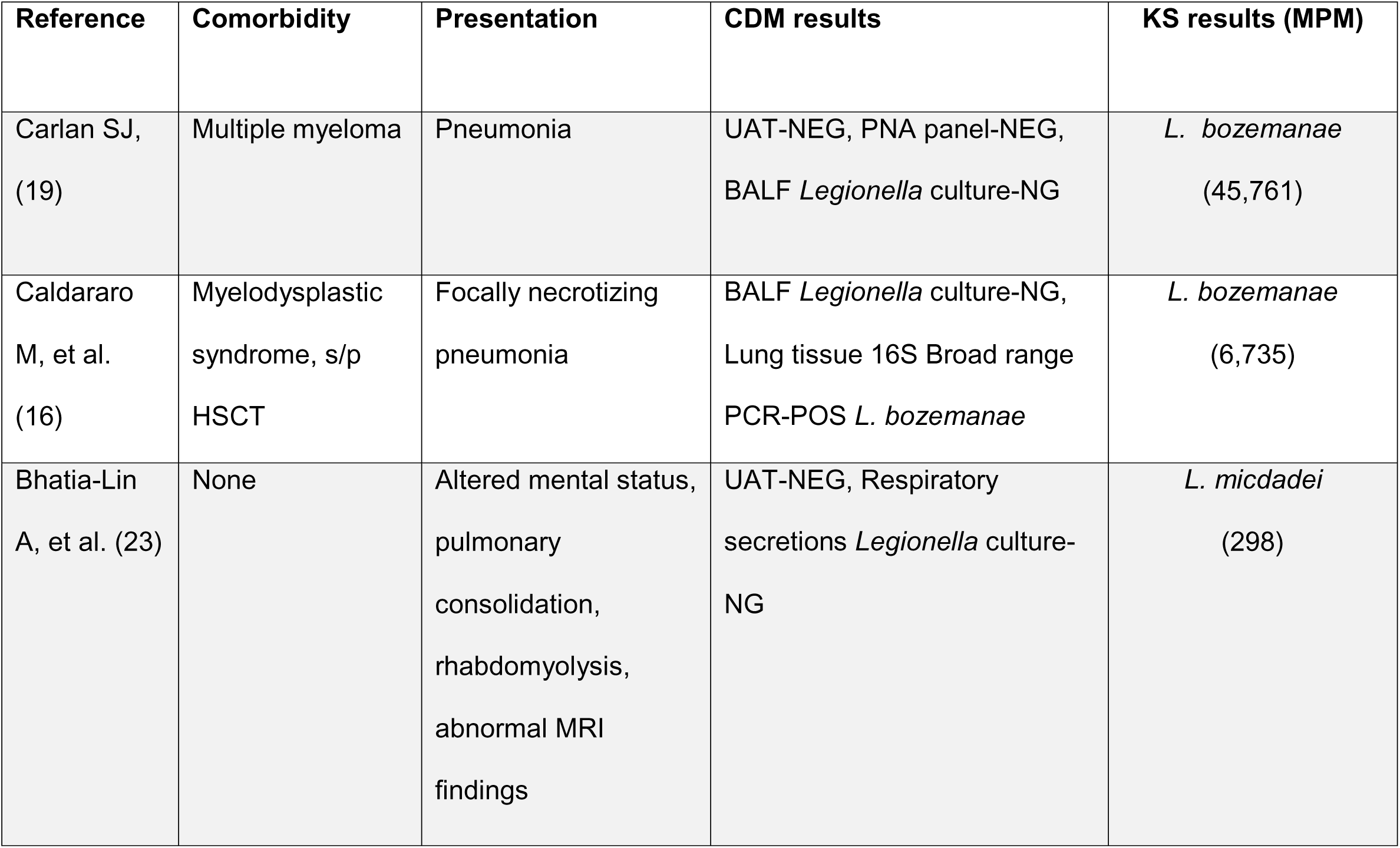
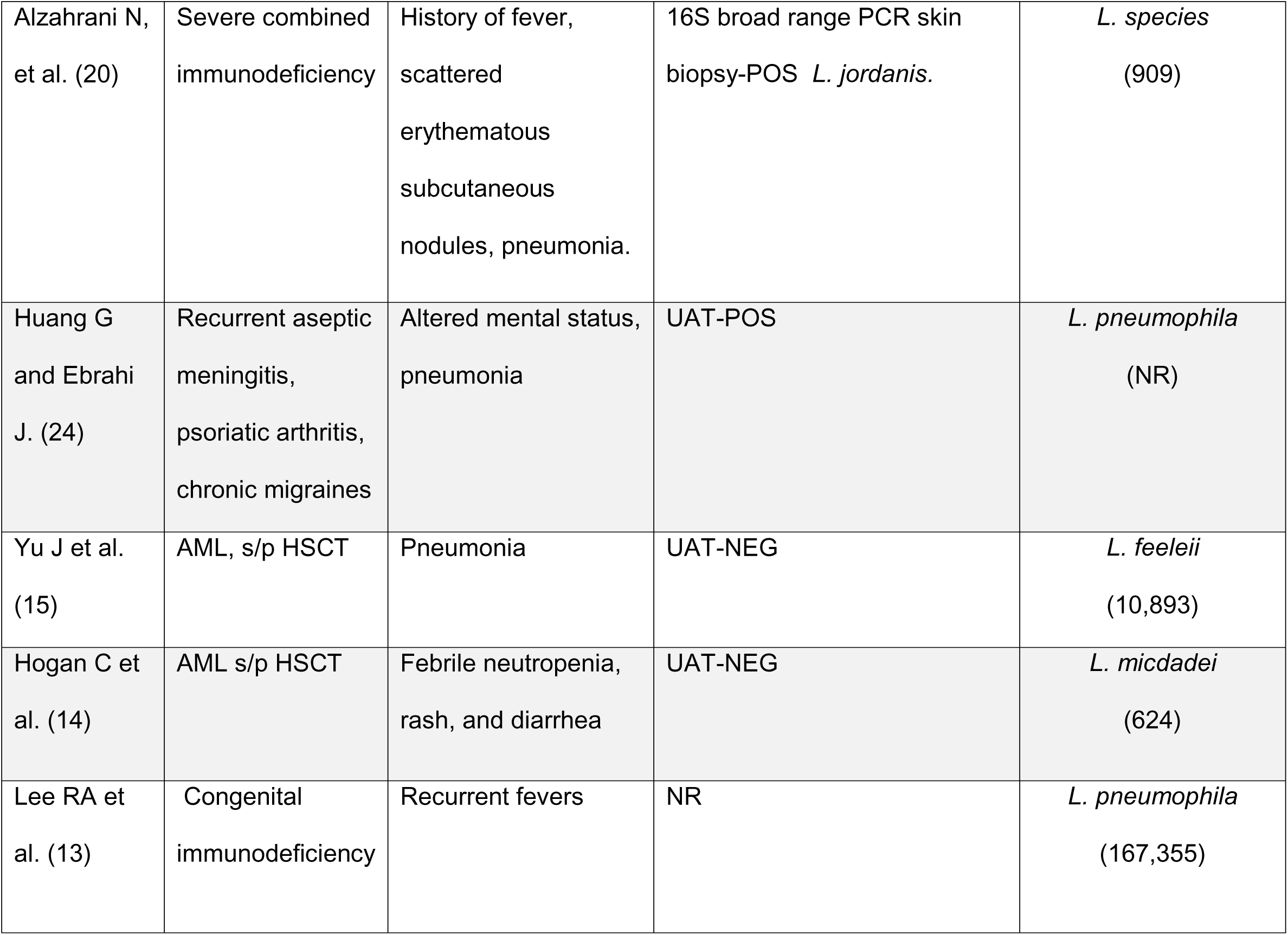
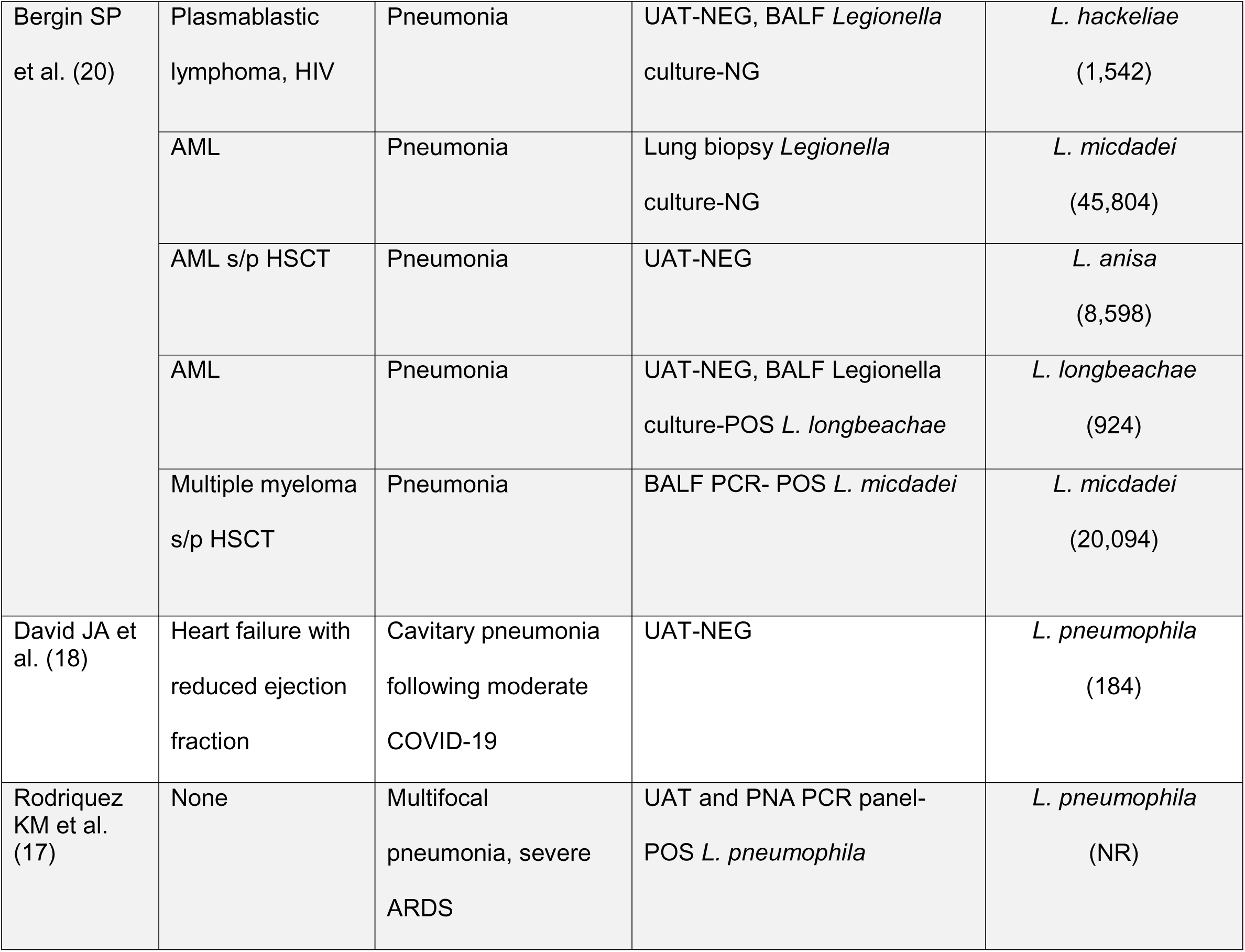
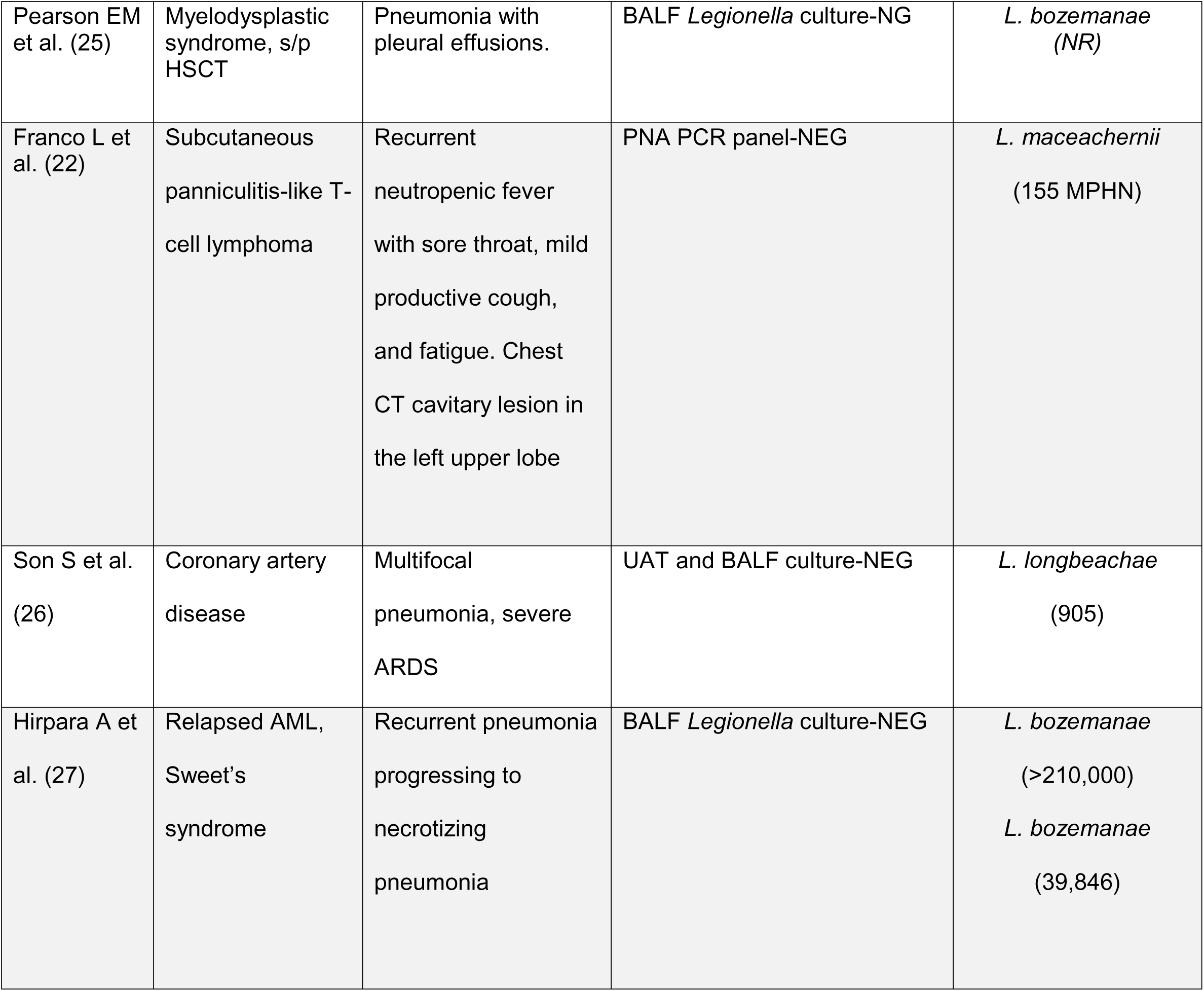
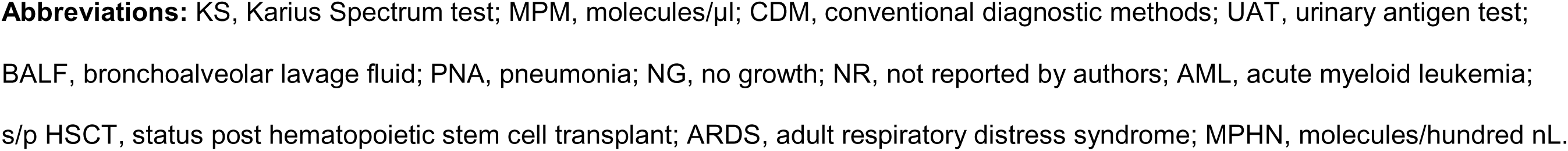
Summary of published reports of plasma mcfDNA sequencing used in the diagnosis of legionellosis.

| Reference | Comorbidity | Presentation | CDM results | KS results (MPM) |
| --- | --- | --- | --- | --- |
| Carlan SJ,<br>(19) | Multiple myeloma | Pneumonia | UAT-NEG, PNA panel-NEG,<br>BALF *Legionella* culture-NG | *L. bozeman*<br>(45,761) |
| Caldararo<br>M, et al.<br>(16) | Myelodysplastic<br>syndrome, s/p<br>HSCT | Focally necrotizing<br>pneumonia | BALF *Legionella* culture-NG,<br>Lung tissue 16S Broad range<br>PCR-POS *L. bozeman* | *L. bozeman*<br>(6,735) |
| Bhatia-Lin<br>A, et al. (23) | None | Altered mental status,<br>pulmonary<br>consolidation,<br>rhabdomyolysis,<br>abnormal MRI<br>findings | UAT-NEG, Respiratory<br>secretions *Legionella* culture-NG | *L. micdadei*<br>(298) |
| Alzahrani N, et al. (20) | Severe combined immunodeficiency | History of fever, scattered erythematous subcutaneous nodules, pneumonia. | 16S broad range PCR skin biopsy-POS *L. jordanis*. | *L. species*<br>(909) |
| Huang G and Ebrahi J. (24) | Recurrent aseptic meningitis, psoriatic arthritis, chronic migraines | Altered mental status, pneumonia | UAT-POS | *L. pneumophila*<br>(NR) |
| Yu J et al. (15) | AML, s/p HSCT | Pneumonia | UAT-NEG | *L. feeleeii*<br>(10,893) |
| Hogan C et al. (14) | AML s/p HSCT | Febrile neutropenia, rash, and diarrhea | UAT-NEG | *L. micdadei*<br>(624) |
| Lee RA et al. (13) | Congenital immunodeficiency | Recurrent fevers | NR | *L. pneumophila*<br>(167,355) |
| Bergin SP<br>et al. (20) | Plasmablastic<br>lymphoma, HIV | Pneumonia | UAT-NEG, BALF *Legionella*<br>culture-NG | *L. hackeliae*<br>(1,542) |
|  | AML | Pneumonia | Lung biopsy *Legionella*<br>culture-NG | *L. micdadei*<br>(45,804) |
|  | AML s/p HSCT | Pneumonia | UAT-NEG | *L. anisa*<br>(8,598) |
|  | AML | Pneumonia | UAT-NEG, BALF *Legionella*<br>culture-POS *L. longbeachae* | *L. longbeachae*<br>(924) |
|  | Multiple myeloma<br>s/p HSCT | Pneumonia | BALF PCR- POS *L. micdadei* | *L. micdadei*<br>(20,094) |
| David JA et<br>al. (18) | Heart failure with<br>reduced ejection<br>fraction | Cavitary pneumonia<br>following moderate<br>COVID-19 | UAT-NEG | *L. pneumophila*<br>(184) |
| Rodriguez<br>KM et al.<br>(17) | None | Multifocal<br>pneumonia, severe<br>ARDS | UAT and PNA PCR panel-POS *L. pneumophila* | *L. pneumophila*<br>(NR) |
| Pearson EM et al. (25) | Myelodysplastic syndrome, s/p HSCT | Pneumonia with pleural effusions. | BALF *Legionella* culture-NG | *L. bozeman*<br>(NR) |
| Franco L et al. (22) | Subcutaneous panniculitis-like T-cell lymphoma | Recurrent neutropenic fever with sore throat, mild productive cough, and fatigue. Chest CT cavitory lesion in the left upper lobe | PNA PCR panel-NEG | *L. maceachernii*<br>(155 MPH) |
| Son S et al. (26) | Coronary artery disease | Multifocal pneumonia, severe ARDS | UAT and BALF culture-NEG | *L. longbeachae*<br>(905) |
| Hirpara A et al. (27) | Relapsed AML, Sweet's syndrome | Recurrent pneumonia progressing to necrotizing pneumonia | BALF *Legionella* culture-NEG | *L. bozeman*<br>(>210,000)<br>*L. bozeman*<br>(39,846) |

**Abbreviations:** KS, Karius Spectrum test; MPM, molecules/ μl; CDM, conventional diagnostic methods; UAT, urinary antigen test; BALF, bronchoalveolar lavage fluid; PNA, pneumonia; NG, no growth; NR, not reported by authors; AML, acute myeloid leukemia; s/p HSCT, status post hematopoietic stem cell transplant; ARDS, adult respiratory distress syndrome; MPHN, molecules/hundred nL.

Detections were concordant with CDM in 6 (31.6%) patients. Only 1 (14%) of 7 *Legionella* cultures was positive (*L. longbeachae*). Two (18%) of 11 UAT were positive. Additionally, there was one false-negative result in a patient where *L. pneumophila* was detected by KS. Molecular methods were used to evaluate 5 (26%) patients. The BioFire Pneumonia Panel was used in 2 patients and was positive in 1 with *L. pneumophila* and negative in 1 with *L. bozemanae* detected by KS. Broad range bacterial PCR and sequencing of the 16S rRNA gene was used for 3 patients. The results were concordant for *L*. *micdadei* and *L. bozemanae,* and discordant in one with *L. jordanis* identified by broad range PCR and as an unresolved *Legionella* sp. identified by DC-3.16. BLAST alignments of mcfDNA sequences in this sample to all assemblies of *Legionella* spp. in the Karius clinical database revealed some relationship to *L. jordanis* above that to the majority of other *Legionella* spp., but far less than that to *L. hackeliae* and species closely related to it (Supplemental Fig. 1B). Analysis of this sample with DC-3.16 strongly supports the call of *Legionella* sp., indicating a species absent from the Karius database. The discrepancy may be due to lack of diversity in the 16S rRNA gene database used, limiting species resolution.

### Hospital A cohort

The KS production results yielded 36 detections of *Legionella* spp. from 34 patients. The calls were 18 (48.6%) *L. pneumophila,* 13 (40.5%) NPLS and 3 (10.8%) unresolved *Legionella* sp. (Fig. 4). The numbers of the nine individual NPLS detected are shown in Fig. 5. Both the relative proportions and spectrum of *Legionella* spp. reflect the larger KS cohort. No patient had multiple species detected in the same sample.

**Figure 4.**
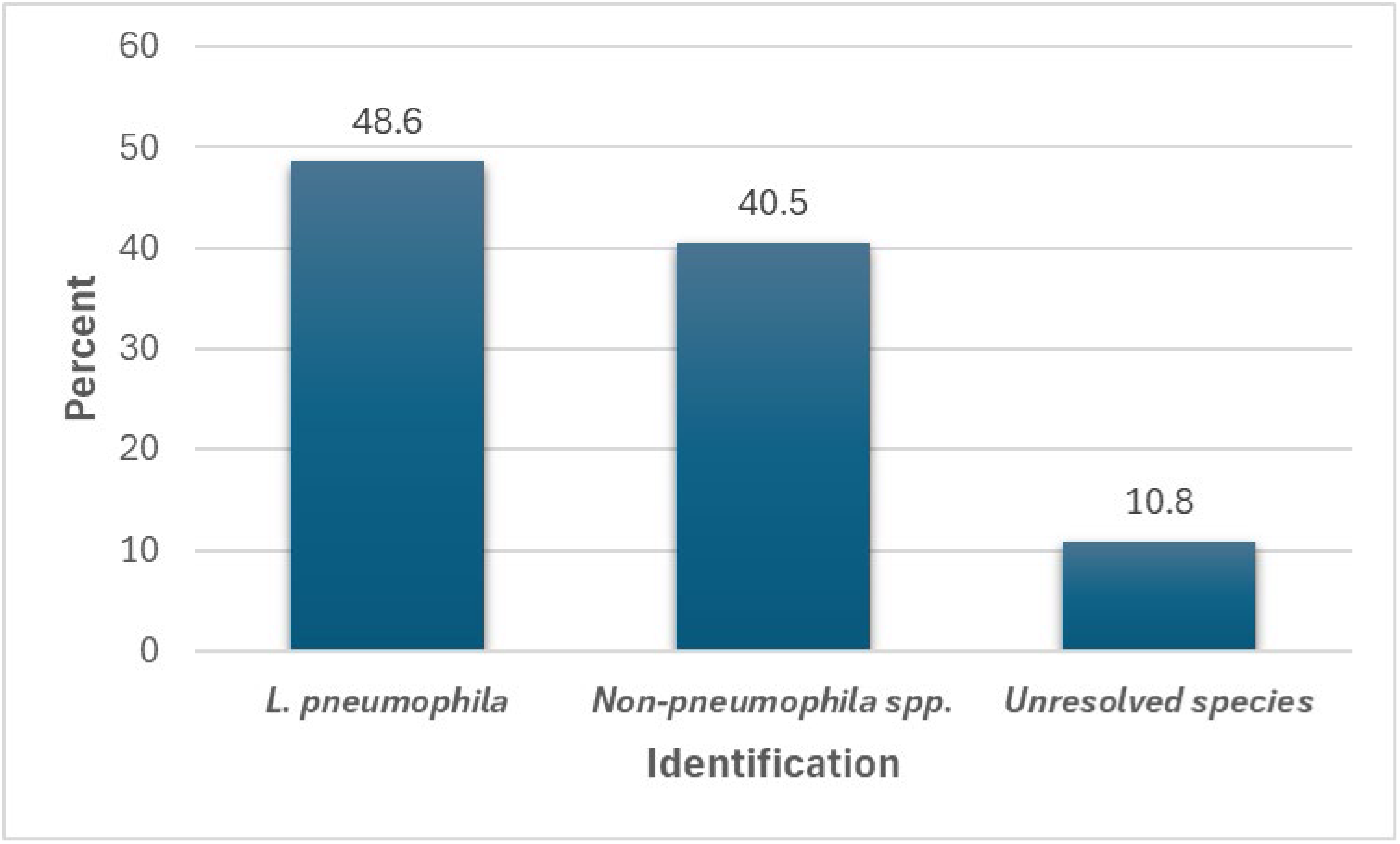
Proportions of *L. pneumophila*, non-*pneumophila* species and unresolved species identified by plasma mcfDNA sequencing in the Hospital A cohort.

**Figure 5.**
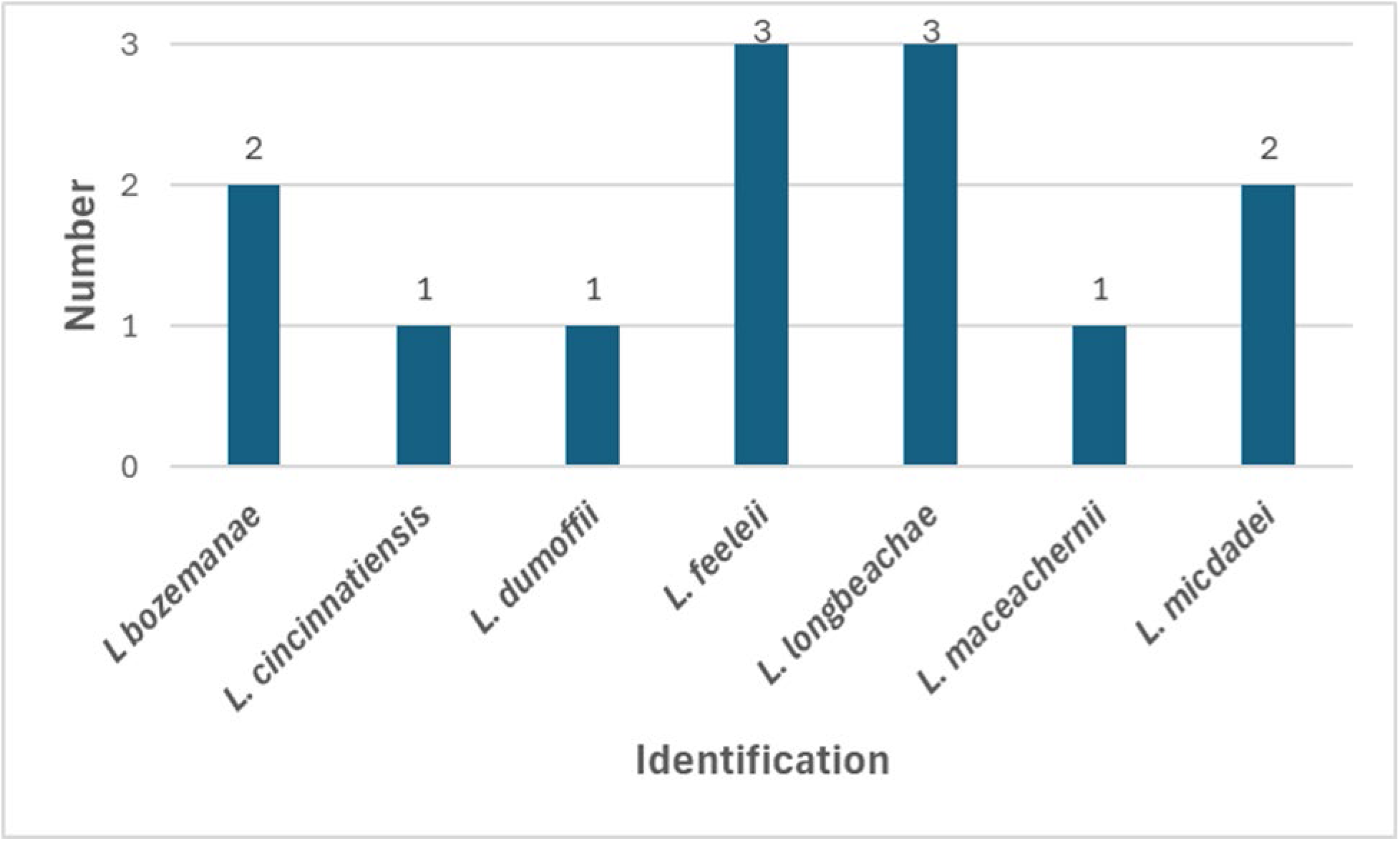
Number of non-*pneumophila* species identified by plasma mcfDNA sequencing in the Hospital A cohort.

The characteristics of the patients included in this cohort are shown in Table 2. Notably, 75.7% were community acquired infections with 91.7% presenting as LD. Two of the patients who presented as LD had extrapulmonary manifestations both with suspected meningitis. A variety of risk factors and comorbidities were noted with 61.8% categorized as immunocompromised patients. Most patients (64.7%) received empiric therapy directed against *Legionella* spp. prior to the report of the KS result. Attributable mortality in this cohort was 8.1%. UAT, culture and PCR were ordered in 91.2%, 8.1% and 2.9%, respectively. UAT was positive in 20.6%, culture was positive for *L. feelei* in one (33.3%) and the single PCR test (ARUP Laboratories, Salt Lake City, Utah) was negative for both *L. pneumophila* and *Legionell*a spp. The diagnosis of legionellosis was made by CDM alone in none, the combination of CDM and KS in 8 (23.5%) and by KS alone in 26 (76.5%). Accordingly, the additive diagnostic value of plasma mcfDNA sequencing was 56.8%. The performance characteristics of UAT for identification of *L. pneumophila* are shown in Table 3. The sensitivity, specificity, and accuracy of the UAT for diagnosis of *L. pneumophila* infections were 46.7% (21.27% to 73.41%), 100% (79.41% to 100.00%), 74.19% (55.39% to 88.14%), respectively. mcfDNA sequencing does not resolve detections of *L. pneumophila* to the serogroup level so it is possible that the 8 false negative UAT occurred in patients with non-serogroup 1 infections. The mean MPM values associated with a positive and negative test were 131,301 and 1,089. Although there was a trend of higher microbial burdens found in patients with a positive UAT the difference in mean values was not significant (*p*=0.2372). The days from hospital admission to collection of KS plasma samples ranged from 0-55 with a median of 3 (IQR 1-6). The days from sample collection to report of results ranged from 1-7 with a median of 2 (IQR 2-3). The KS test results provided a new diagnosis in 72.9%, an earlier diagnosis in 5.4%, and confirmed the diagnosis in 16.2% of patients. The results led to directed therapy for legionellosis in 29.7%.

**Table 2.** Characteristics of the 34 patients with plasma mcfDNA detections of *Legionella* spp, at Hospital A.

| **Characteristics** |  |
| --- | --- |
| **Age, Years** |  |
| Median (IQR) | 57 (44-66) |
| Range | 17-80 |
| **Male sex, No. (%)** | 21 (61.8%) |
| **Acquisition, No. (%)** |  |
| Community | 25 (73.5) |
| Health care facility | 5 (14.7) |
| Not reported | 4 (11.8) |
| **Presentation, No. (%)** |  |
| Legionnaires' disease | 33 (97.1) |
| Not reported | 1(2.9) |
| **Risk factors, No. (%)** |  |
| None | 7 (20.6) |
| SOT | 6 (17.6) |
| BMT | 5 (14.7) |
| Immune system disorder, nos | 2 (5.9) |
| AML on CTx | 2 (5.9) |
| Obesity | 2 (5.9) |
| SLE on IST | 2 (5.9) |
| Smoking (historic) | 1 (2.9) |
| APL not on CTx | 1 (2.9) |
| Asplenia | 1 (2.9) |
| Chronic Lung Disease | 1 (2.9) |
| Hepatic failure | 1 (2.9) |
| Malignancy nos | 1 (2.9) |
| MDS on CTx | 1 (2.9) |
| Rheumatoid arthritis on IST | 1 (2.9) |
| **Targeted Tx prior to results, No., (%)** | 22 (64.7) |
| **Other *Legionella* tests, No. tested, (% positive)** |  |
| UAT | 31 (22.6) |
| Culture | 3 (33.3) |
| PCR | 1 (0) |
| **Mortality, No. (%)** | **3\* (8.8)** |

\*1 expired due to liver failure
**Abbreviations:** SOT, solid organ transplant; BMT, bone marrow transplant; nos, not otherwise specified; AML, acute myeloid leukemia; CTx, chemotherapy; SLE, systemic lupus erythematosus; IST, immunosuppressive therapy; MDS, myelodysplastic syndrome; Tx, treatment; UAT, urinary antigen test.

**Table 3.** Performance characteristics of the urinary antigen test for identification of *L. pneumophila* at Hospital A.

| UAT | ***Legionella* Identifications by mcfDNA sequencing** |  |  |
| --- | --- | --- | --- |
|  | *L. pneumophila* | Other species | Sum |
| Positive | 7 | 0 | 7 |
| Negative | 8 | 16 | 24 |
| Sum | 15 | 16 | 31 |

## DISCUSSION

Accurate diagnosis of legionellosis depends on clinical suspicion and appropriate laboratory testing. However, current diagnostic strategies disproportionately rely on UAT, which detects only Lp1 and likely underestimates infections caused by other species (28). Culture and PCR, although capable of broader detection, are inconsistently performed and may fail to detect certain NPLS. These diagnostic constraints shape both patient-level management and national surveillance data.

In this multi-cohort analysis, plasma mcfDNA sequencing identified a substantially higher proportion of NPLS compared with CDC confirmed cases. The reduction in unresolved species calls following application of the DC-3.16 phylogeny-based filter further supports improved species-level resolution. Together, these findings suggest that the predominance of *L. pneumophila* in surveillance data may reflect diagnostic bias rather than true epidemiology.

Contamination of molecular reagents with environmental *Legionella* DNA is a recognized concern (29). In this study, low-level background reads were detected in a small fraction of NTC, but reporting thresholds and batch-level review procedures effectively mitigated the risk of false-positive results. Similarly, low-level detections of *L. maceachernii* in asymptomatic donors rarely met reporting criteria. These findings support the analytical robustness of the platform under real-world conditions. It should be noted that colonization with *L. maceachernii* has not been previously described. However, colonization with *L. pneumophila* has been rarely reported (30, 31).

In the patients with *Legionella* spp. detected in multiple samples the same species was detected in each patient supporting the clinical reproducibility of the results. The microbial burden kinetics and long duration mcfDNA signal in these patients are more difficult to interpret in the absence of clinical context. However, 85% showed a significant decrease in signal over time consistent with response to treatment and the median duration of signal of 24.5 days provides a potentially wider diagnostic window than culture since viable bacteria are rapidly eliminated by antibiotics. The literature review and the Hospital A cohort underscore the clinical relevance of broader species detection. Published cases primarily involved immunocompromised patients, frequently required invasive sampling, and were often attributable to NPLS In these reports, concordance with conventional diagnostics was limited. In the Hospital A cohort, UAT was widely ordered but culture and PCR were infrequently performed. Plasma mcfDNA sequencing provided a new or earlier diagnosis in most cases and demonstrated substantial additive diagnostic value.

Importantly, most infections would not have been identified by conventional testing alone. Operational metrics further support clinical utility mcfDNA sequencing. Median time from hospital admission to sample collection was short, and results were typically reported within two days of sample receipt, enabling timely diagnostic clarification.

The observed sensitivity of UAT for *L. pneumophila* infections in the Hospital A cohort was limited, consistent with prior literature. Although higher plasma microbial burdens were associated with UAT positivity, this relationship was not statistically significant, suggesting that additional biological or technical factors influence UAT performance. These findings reinforce that UAT should not be relied upon as a standalone diagnostic modality in suspected legionellosis.

At the population level, incorporation of broader molecular diagnostics may help clarify the epidemiology of *Legionella* infections. Surveillance systems built around UAT are inherently biased toward detecting Lp1 infections. While mcfDNA sequencing does not provide strain-level resolution required for outbreak source attribution, earlier and more comprehensive species identification may support timely recognition of unusual patterns and inform targeted investigations.

This study has limitations. Detailed clinical data were unavailable for most patients in the KS cohort, restricting assessment of clinical relevance at scale. The Hospital A analysis, although informative, included a modest number of patients and incomplete comparator testing, precluding definitive sensitivity estimates for either modality. In addition, the KS cohort was enriched for hospitalized and immunocompromised individuals, which may limit generalizability to lower-risk populations.

Despite these limitations, the convergence of large-scale cohort data, published case reports, and real-world institutional experience suggests that plasma mcfDNA sequencing expands detection of NPLS and may improve diagnostic yield in selected high-risk populations. Broader integration of unbiased molecular diagnostics has the potential to refine understanding of disease burden and complement existing surveillance frameworks.

## Data availability

The Karius Test is a proprietary laboratory-developed test, and thus there are several limitations on data availability that we declare here. Karius is unable to share the raw sequencing data underlying the test results evaluated here due to issues relating to patient privacy and consent. Additionally, the methodological details are protected by Karius’s intellectual property and are not available for third party use. Taking these two limitations into account, summarized data used to reach the conclusions in this study are included in the manuscript and supplemental material.

## Supporting information

Data Availability Tables

## ACKNOWLEDGEMENTS

Frederick S. Nolte, Martin S. Lindner, Shivkumar Venkatasubrahmanyam, Sarah Y. Park, Bradley A. Perkins are employees of Karius, the company that markets the commercial test evaluated in this study. These authors contributed to the study design, data analysis, and the writing and editing of the manuscript. Richard G. Wunderink serves as a consultant for Karius and contributed to the writing and editing of the manuscript. Lindsay Lim and Vincent P. Hsu report no conflicts of interest and contributed to data collection and the writing and editing of the manuscript. Chiagozie I. Pickens reports no conflicts of interest and contributed to the writing and editing of the manuscript.

The authors thank the entire Karius team for their expertise, dedication, and collaboration, which made this study possible. This study was solely funded by Karius.

