## Supplemental Table 1 and Figure 1 for "Plasma Microbial Cell-Free DNA Metagenomic Sequencing Bridges Gaps in the Diagnosis, Epidemiology and Surveillance of *Legionella* Infections"

**Supplemental Table 1.** Patients with *Legionella* spp. detected by the Karius Spectrum test in multiple samples.

| **Pt no.** | **Date reported** | **Species** | **MPM^a^** | **Log MPM** | **Duration (days)** | **Log difference^b^** |
| --- | --- | --- | --- | --- | --- | --- |
| 1 | 2019-09-15 | *L. maceachernii* | 1,035 | 3.02 | 433 | 0.89 |
|  | 2020-11-21 | *L. maceachernii* | 7,970 | 3.90 |  |  |
| 2 | 2022-11-12 | *L. jordanis* | 154,649 | 5.19 | 20 | -3.34 |
|  | 2022-12-02 | *L. jordanis* | 71 | 1.85 |  |  |
| 3 | 2023-10-11 | *L. species* | 616,717 | 5.79 | 23 | -2.03 |
|  | 2023-11-03 | *L. species* | 5699 | 3.76 |  |  |
| 4 | 2022-03-25 | *L. bozemanae* | 22,480 | 4.35 | 58 | -2.45 |
|  | 2022-04-10 | *L. bozemanae* | 1,089 | 3.04 |  |  |
|  | 2022-05-04 | *L. species* | 285 | 2.46 |  |  |
|  | 2022-05-22 | *L. bozemanae* | 80 | 1.90 |  |  |
| 5 | 2021-06-15 | *L. maceachernii* | 14,410 | 4.16 | 12 | -0.47 |
|  | 2021-06-27 | *L. maceachernii* | 4,909 | 3.69 |  |  |
| 6 | 2023-05-11 | *L. longbeachae* | 63,081,265 | 7.80 | 14 | -5.37 |
|  | 2023-06-01 | *L. longbeachae* | 270 | 2.43 |  |  |
| 7 | 2023-12-23 | *L. longbeachae* | 64 | 1.81 | 24 | -0.31 |
|  | 2024-01-06 | *L. longbeachae* | 32 | 1.50 |  |  |
| 8 | 2020-05-29 | *L. pneumophila* | 30,401 | 4.48 | 7 | -0.94 |
|  | 2020-06-05 | *L. pneumophila* | 3,483 | 3.54 |  |  |
| 9 | 2024-04-07 | *L. micdadei* | 1,291,351 | 6.11 | 41 | -1.56 |
|  | 2024-04-17 | *L. micdadei* | 521,523 | 5.72 |  |  |
|  | 2024-05-18 | *L. micdadei* | 35,530 | 4.55 |  |  |
| 10 | 2023-12-30 | *L. pneumophila* | 4,289,667 | 6.63 | 39 | -4.10 |
|  | 2024-01-07 | *L. pneumophila* | 2,921,807 | 6.47 |  |  |
|  | 2024-01-19 | *L. pneumophila* | 62753 | 4.80 |  |  |
|  | 2024-01-31 | *L. pneumophila* | 540 | 2.73 |  |  |
|  | 2024-02-07 | *L. pneumophila* | 342 | 2.53 |  |  |
| 11 | 2024-07-18 | *L. micdadei* | 1,996 | 3.30 | 62 | 0.04 |
|  | 2024-09-18 | *L. micdadei* | 2,170 | 3.34 |  |  |
| 12 | 2024-06-27 | *L. species* | 385 | 2.59 | 1 | -0.15 |
|  | 2024-06-28 | *L. species* | 274 | 2.44 |  |  |
| 13 | 2023-10-13 | *L. micdadei* | 33,124 | 4.52 | 49 | -1.90 |
|  | 2023-12-01 | *L. micdadei* | 413 | 2.62 |  |  |
| 14 | 2020-06-24 | *L. species* | 2,840,336 | 6.45 | 78 | -4.39 |
|  | 2020-09-10 | *L. species* | 116 | 2.06 |  |  |
| 15 | 2024-04-28 | *L. pneumophila* | 129,792 | 5.11 | 25 | -2.81 |
|  | 2024-05-23 | *L. pneumophila* | 201 | 2.30 |  |  |
| 16 | 2023-02-26 | *L. pneumophila* | 354 | 2.55 | 11 | -0.31 |
|  | 2023-03-09 | *L. pneumophila* | 174 | 2.24 |  |  |
| 17 | 2021-04-25 | *L. species* | 224,364 | 5.35 | 24 | -2.41 |
|  | 2021-05-06 | *L. species* | 3187 | 3.50 |  |  |
|  | 2021-05-19 | *L. species* | 868 | 2.94 |  |  |
| 18 | 2022-07-07 | *L. bozemanae* | 209,961 | 5.32 | 28 | -3.02 |
|  | 2022-07-16 | *L. bozemanae* | 7,493 | 3.87 |  |  |
|  | 2022-07-28 | *L. bozemanae* | 684 | 2.83 |  |  |
|  | 2022-07-31 | *L. bozemanae* | 1,621 | 3.21 |  |  |
|  | 2022-08-04 | *L. species* | 199 | 2.30 |  |  |
| 19 | 2023-05-11 | *L. pneumophila* | 677 | 2.83 | 78 | 4.33 |
|  | 2023-07-28 | *L. pneumophila* | 14,419,691 | 7.16 |  |  |
| 20 | 2023-02-26 | *L. pneumophila* | 5,132 | 3.71 | 11 | -1.16 |
|  | 2023-03-09 | *L. pneumophila* | 354 | 2.55 |  |  |

^a^Molecules/µl.

^b^Between first and last sample.

**A.**

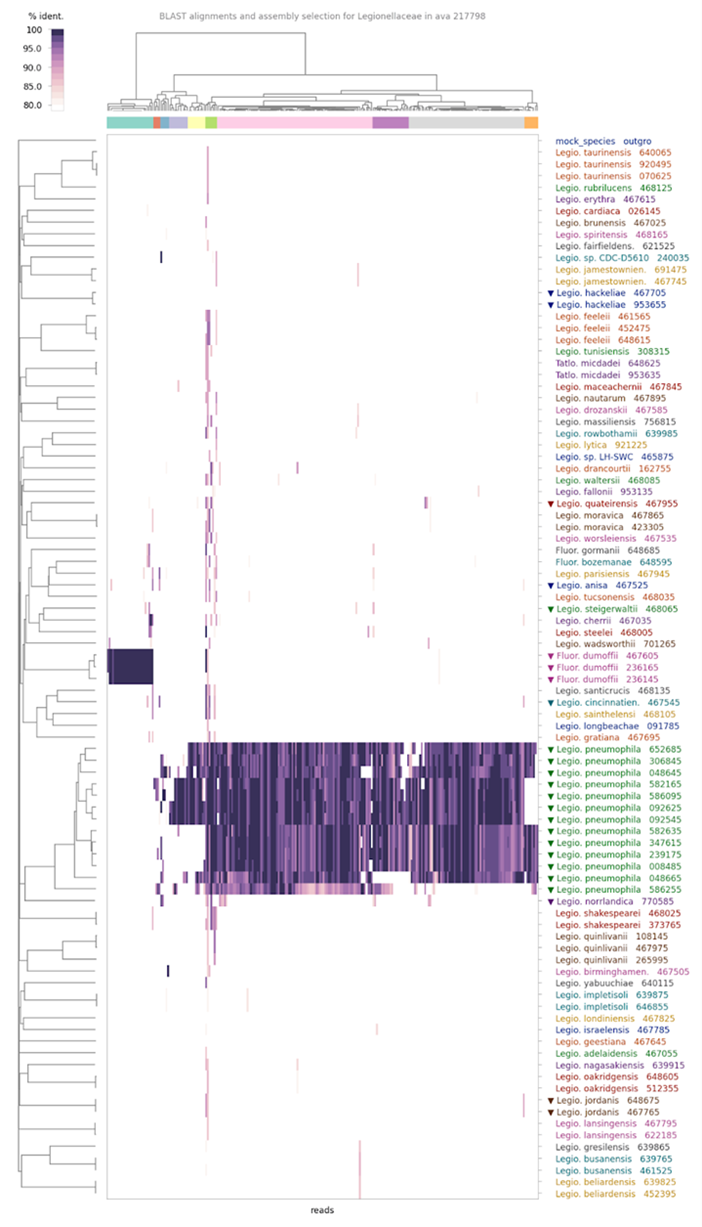

**B.**

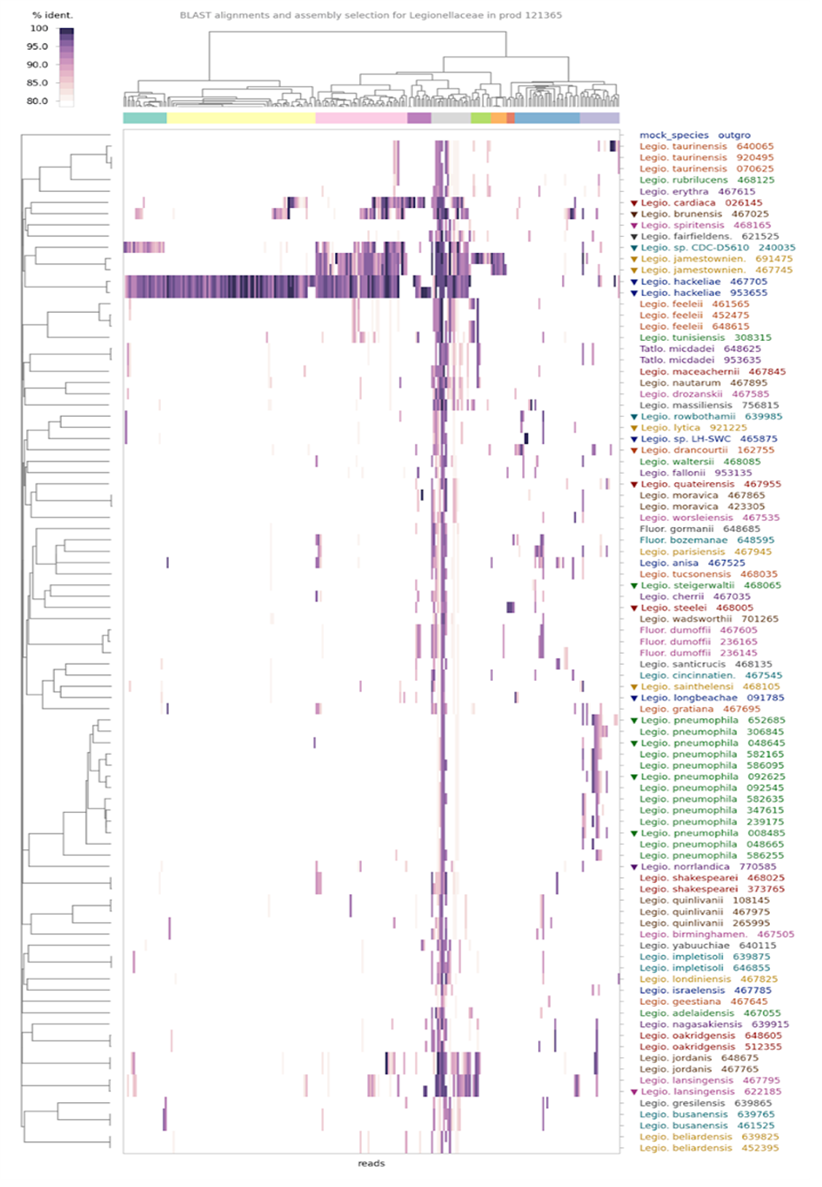

**Supplemental Figure 1.** Figure S1: BLAST alignment of mcfDNA sequences supports species identification by pipeline DC-3.16. In each panel, the heatmap shows BLAST percent identity (dark purple indicates a perfect match) of a random sample of 250 mcfDNA fragments (columns) to whole genome assemblies (rows) in the family *Legionellaceae* that are present in the Karius database. The left margin shows a whole genome phylogenomic tree. The upper margin shows hierarchical clustering of reads. (A) In this sample, 3.16 identified a mixture of around 90% *L. pneumophila* and 10% *L. dumoffii.* This is supported by the presence of non-overlapping sets of mcfDNA fragments of these proportions, which align to each of these species. (B) Only a small fraction of mcfDNA sequences align to *L. jordanis*. These and most other fragments align to *L. hackeliae,* and to a lesser extent, *L. jamestownensis, L. clemsonensis* and *L. cardiaca*. However, most of these alignments are imperfect, and these species do not account for the remaining 25-30% of fragments. Together, this pattern is inconsistent with *L. jordanis* identified by broad-range PCR, but consistent with the DC-3.16 identification of “*Legionella* species”, indicating a species absent from the Karius database.
